# Bubbles help in troubles: contrast-enhanced ultrasound (CEUS) as predictor of recurrence for TIA/stroke in low-grade internal carotid artery stenosis

**DOI:** 10.1101/2024.04.17.24306003

**Authors:** Francesca D’Auria, Danilo Flavio Santo

## Abstract

**Introduction:** Contrast-enhanced ultrasound (CEUS) allows to visualize atherosclerotic plaque neo-vessels which are the hallmark of carotid plaque instability.

**Aim:** Aim of our prospective study was to check the correlation between carotid CEUS analysis and the recurrence of TIA/stroke in patients with a previous recent TIA/stroke and neurological impairment congruent with the vascular stenosis.

**Materials and Methods:** From November 2021 to May 2023, 62 consecutive patients (mean age 73.8 +/- 12.2, female 51) with a TIA/stroke in the previous 30 days, underwent carotid ultrasound and carotid CEUS in outpatients setting after 10 days from the acute event. Inclusion criteria was one atherosclerotic plaque inside the internal carotid artery, congruent with symptoms, which was causing a stenosis less than 50% (low-grade stenosis). Carotid plaque neovascularization scoring method was: score 0: no visible microbubbles within the plaque (A); score 1: minimal microbubbles confined to periadventitial (B); and score 2: microbubbles present throughout the plaque core (C). During the six-month follow-up, we checked TIA/stroke recurrences. A multivariable logistic regression analysis was performed.

**Results:** In our series, 22% of patients have CEUS score 0, 35% CEUS score 1, and 43% CEUS score 2. At six-month follow-up we found 21% TIA/stroke recurrences in CEUS score 2, despite of the ongoing best medical therapy as per guidelines. In Cox regression analysis, CEUS-detected neovascularization was independently associated with TIA/stroke recurrence (hazard ratio, 5.37; 95% confidence interval, 1.36–2.31).

**Conclusions:** Plaque neovascularization, detected by CEUS, is an independent predictor of TIA/stroke recurrence at six-month follow-up in patients with carotid atherosclerosis despite of low-grade stenosis.

## Introduction

International guidelines for carotid artery atherosclerosis surgical and/or endovascular treatment are clear and strictly related to patients’ symptoms, the grade of vascular stenosis, and the measured velocity^1-2^. In case of not significant stenosis caused by soft plaque which may determine TIA/stroke and its recurrence due to embolism, surgeons are keen to remove it^3^. This prudential orientation may cause an overtreatment of patients. In regards of this perceived feeling of surgical overdo and aiming to move towards increasingly precise medicine, we started a prospective observational trial with the purpose to identify possible diagnostic strategy to better orient surgical decision making in this grey zone. To meet our commitment, reducing not only surgical overtreatment but also patients X-ray exposure, we find that Carotid Contrast-Enhanced Ultrasound (CEUS) may be the methods to adopt in our research program and we try to elaborate an image-based surgical decision score system. In fact, CEUS is an imaging technique that involves the use of contrast agents (e.g. saline solution bubble or sulphur hexafluoride solution) to enhance the visualization of blood flow within the carotid arteries using ultrasound technology. During a carotid CEUS procedure, the contrast agent is injected into the bloodstream. It highlights the blood vessels and improves the clarity of the ultrasound images. This allows to more accurately evaluate the structure and function of the carotid arteries, aiding in the diagnosis and management of conditions such as carotid artery disease and stroke risk assessment^4-5^.

## Material and Methods

Aim of our prospective study was to check the correlation between carotid CEUS analysis and the recurrence of TIA/stroke in patients with a previous recent TIA/stroke and neurological impairment congruent with the vascular stenosis. From November 2021 to February 2023, 62 consecutive patients (mean age 73.8 +/- 12.2, female 51) with a TIA/stroke in the previous 30 days, underwent carotid ultrasound and carotid CEUS using intravenous 3 ml sulphur hexafluoride solution (SonoVue^®^) in outpatients setting after 10 days from the acute event. The demography of the population is summarized in Table 1. Inclusion criteria was one atherosclerotic plaque inside the internal carotid artery, congruent with symptoms, which was causing a stenosis less than 50% (low-grade stenosis), while exclusions criteria were history of cerebral vascular diseases; history of neurological disease; presence of infections, malignant tumours, cardiopulmonary dysfunction, hepatic dysfunction, kidney failure (any degree), respiratory failure; hyperechoic or uniformly hyperechoic plaque. We arbitrary established a carotid plaque neovascularization scoring method as it is presented in Figure 1. The score 0 means no visible microbubbles within the plaque (A); score 1 is when minimal microbubbles are confined to peri adventitial (B); and score 2 consists in microbubbles which are present throughout the plaque core (C). During the six-month we checked TIA/stroke recurrences. No one patient was lost at follow-up. A multivariable logistic regression analysis for ischemic stroke and recurrent TIA was performed checking correlation between TIA/stroke recurrence and gender, age, hypertension, hyperlipidaemia, diabetes (any type), chronic obstructive lung disease (COPD), smoke status (active or previous), obesity, stress, atrial fibrillation, plaque velocity (peak systolic velocity), internal carotid kinking, CEUS score 0, CEUS score 1, and CEUS score 2. Internal ethical committee approved the study and produced a specific informed consent form for patients. These study compliances with ethical standards; each patient signed the specific informed consent (in accordance with the Declaration of Helsinki) to be enrolled in this research and authorized to publish the data in anonymous mode for search purpose only. An additional informed consent for personal data collection, treatment, post processing analysis, and publishing purposes was collected and signed by each patient as per institutional internal ethical committee. No sponsor or external fund were used to conduct this independent search. Statistical analysis was performed using SPSS Statistics 17.0 software. Frequency and percentages (%) were used for categorical variables, whereas mean ± SD were employed for continuous variables. Continuous variables were compared between three groups using the independent t test. Proportions were compared using the chi-square test. To determine the association between variables and ischemic stroke or recurrent TIA after TIA, a multivariate logistic regression analysis was performed. Receiver operation curve (ROC) was generated to examine the efficacy of the resulting model. The level of significance was set at P value < 0.05.

**Table 1.** Patients’ demography.

| *Variable* | *N (%) / Mean ± SD* | *Variable* | *N (%) / Mean ± SD* |
| --- | --- | --- | --- |
| Female | 51 (82) | Antiplatelet therapy | 60 (97) |
| Male | 11 (18) | Antihypertensive therapy | 62 (97) |
| Age | 73.8 +/- 12.2 | Statin | 58 (94) |
| Hypertension | 60 (97) | New Oral Anticoagulant | 22 (35) |
| Hyperlipidaemia | 58 (94) | Atrial fibrillation | 8 (13) |
| Diabetes (any type) | 24 (39) | Coronary artery disease | 2 (3) |
| COPD | 21 (34) | Peripheral artery disease | 14 (23) |
| Active smoker | 41 (66) | Internal carotid artery stenosis (%) | 52 +/- 10 |
| Previous smoker | 21 (34) | Peak systolic velocity (cm/s) | 162 +/- 104 |
| Stress | 32 (52) | End diastolic velocity (cm/s) | 62 +/- 12 |

**Figure 1.**
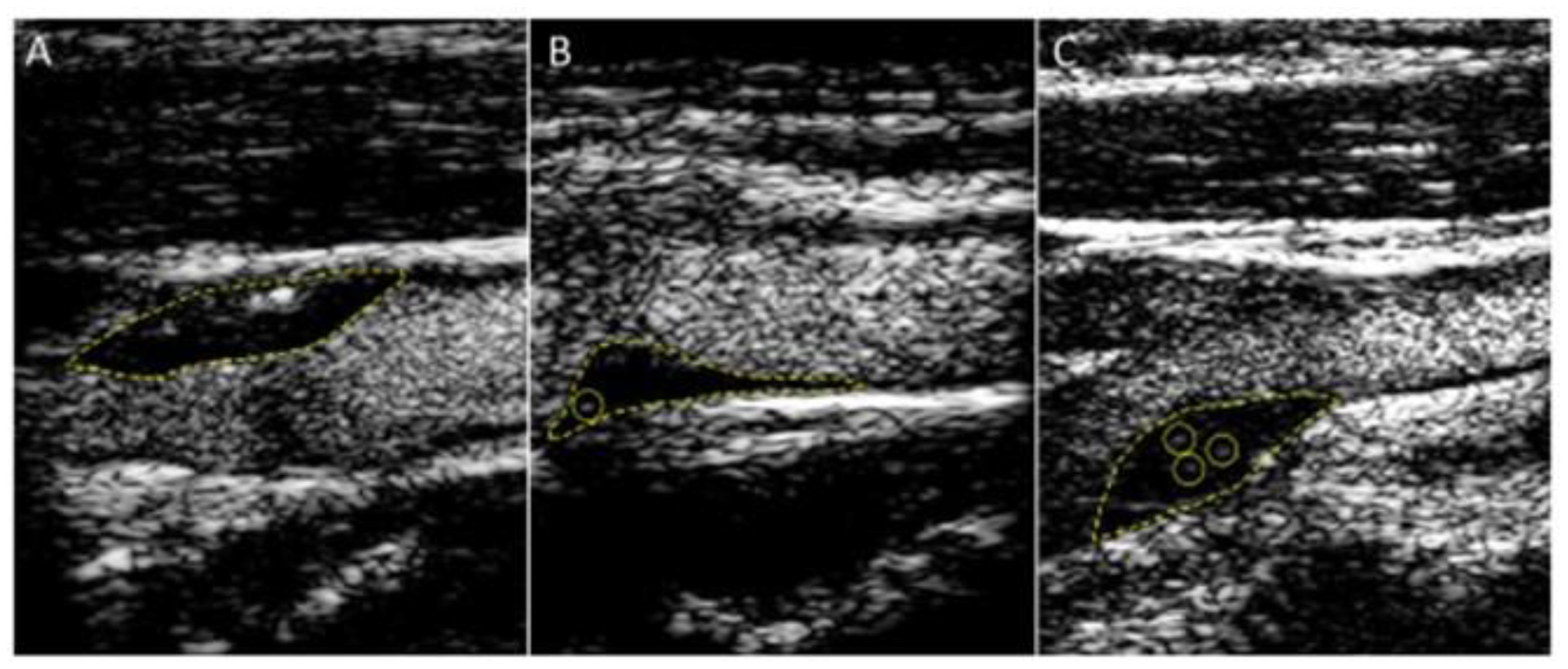
Carotid plaque neovascularization scoring: score 0: no visible microbubbles within the plaque (A); score 1: minimal microbubbles confined to periadventitial (B); and score 2: microbubbles present throughout the plaque core (C).

## Results

In our series, 22% of patients have a CEUS score 0; 35% of patients have a CEUS score 1; and 43% of patients have a CEUS score 2. At six-month follow-up we found 21% TIA/stroke recurrences in CEUS score 2, despite of the ongoing best medical therapy as per guidelines.

The multivariable logistic regression analysis for ischemic stroke and recurrent TIA, shows that CEUS-detected neovascularization was independently associated with TIA/stroke recurrence (hazard ratio, 5.37; 95% confidence interval, 1.36–2.31), while no other considered factors appeared to be significant as predictors of recurrence TIA/stroke, as it is reported in Table 2.

**Table 2.** Multivariable logistic regression analysis for ischemic stroke/TIA recurrence.

| *Variable* | *P value* | *Hazard Ratio* | *Confidence Interval 95%* |
| --- | --- | --- | --- |
| Female | 0.21 | 0.07 | 0.67 – 0.87 |
| Male | 0.11 | 0.04 | 0.57 – 0.98 |
| Age | 0.89 | 0.02 | 0.76 – 0.89 |
| Hypertension | 0.98 | 0.32 | 0.57 – 0.98 |
| Hyperlipidaemia | 0.92 | 1.2 | 0.26 – 0.29 |
| Diabetes (any type) | 0.76 | 1.8 | 0.76 – 0.89 |
| COPD | 0.09 | 0.5 | 0.06 – 0.89 |
| Active smoker | 0.78 | 0.7 | 0.23 – 9.87 |
| Previous smoker | 0.43 | 0.3 | 0.57 - 12.48 |
| Obesity | 0.08 | 1.3 | 0.05 - 11.28 |
| Stress | 0.68 | 0.2 | 0.26 – 0.98 |
| Atrial fibrillation | 0.89 | 1.8 | 0.76 – 0.89 |
| Peak systolic velocity | 0.25 | 1.98 | 0.22 – 0.34 |
| Vessel kinking | 0.82 | 1.72 | 0.52 - 9.48 |
| CEUS score 0 | 0.23 | 1.02 | 0.21 – 1.28 |
| CEUS score 1 | 0.08 | 1.32 | 1.28-1.98 |
| **CEUS score 2** | **0.002** | **5.37** | **1.36–2.31** |

The ROC of the multivariate logistic regression analysis model for CEUS 2 score is showed in Figure 2.

**Figure 2.**
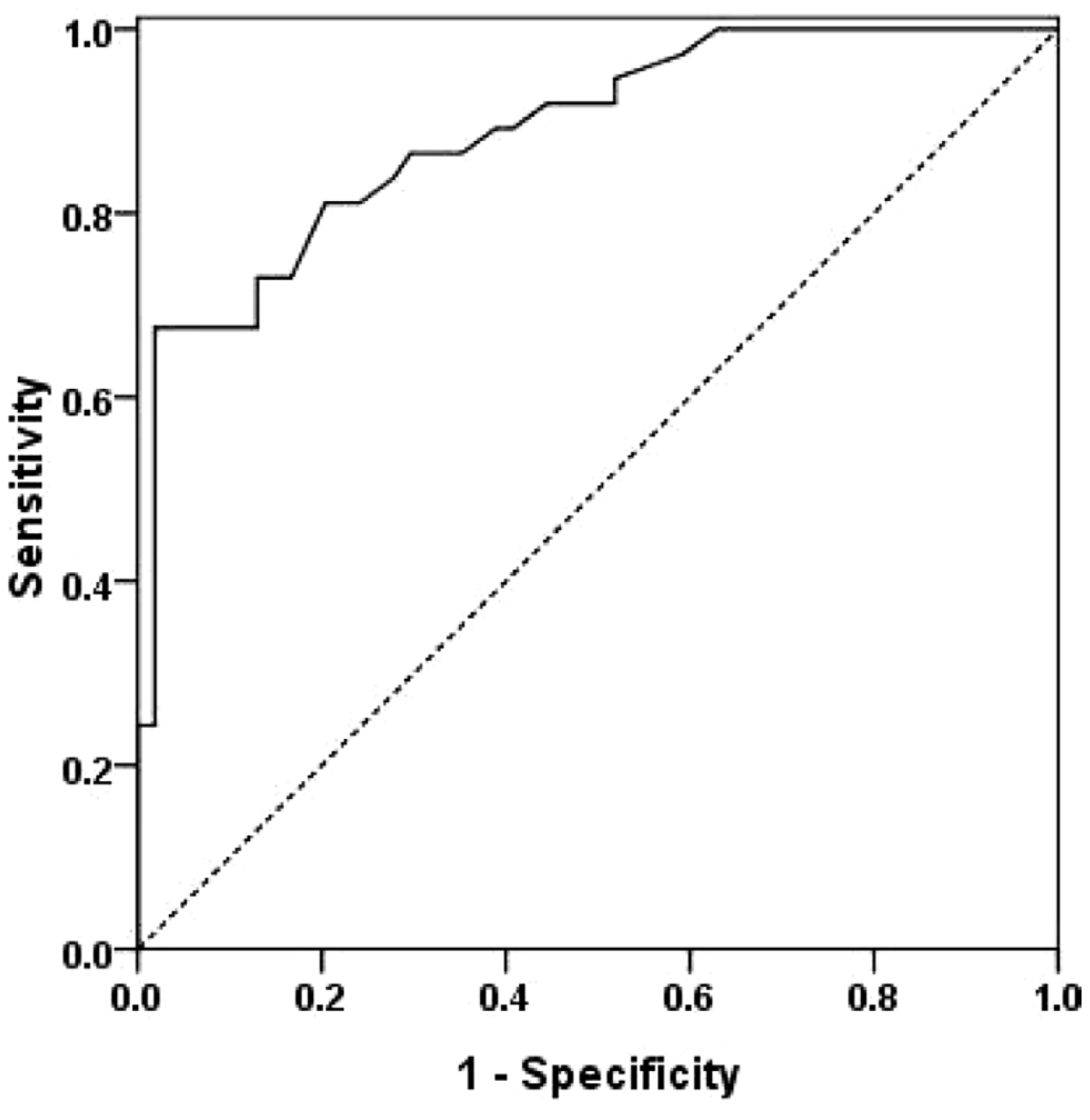
Receiver operation curve (ROC) of the multivariate logistic regression analysis model (CEUS 1, CEUS 2 score).

## Discussion

Carotid CEUS is often used as a non-invasive alternative to other imaging techniques such as computed tomography angiography (CTA) and/or magnetic resonance angiography (MRA) for evaluating extra-cranial carotid artery disease^6^. It offers advantages such as real-time imaging, lack of ionizing radiation, and the ability to assess blood flow dynamics. Carotid plaque instability refers to a condition where the atherosclerotic plaque becomes particularly prone to rupture or erosion. When plaque becomes unstable, it can lead to the formation of blood clots (thrombosis) or embolism. These clots or particles can then travel to smaller blood vessels in the brain, leading to blockages and potentially causing a stroke or transient ischemic attack (TIA)^3^. Several factors contribute to carotid plaque instability, including the composition of the plaque because plaque that contains a large amount of soft, lipid-rich material is more prone to rupture than stable, calcified plaque. Other factors for instability are the plaque size and shape; in fact, large, irregularly shaped plaques are more likely to be unstable compared to smaller, smoother plaques. Underlying medical conditions like hypertension, diabetes, hyperlipidaemia, and smoking can contribute to plaque instability by promoting inflammation and atherosclerosis; finally, hemodynamic factors, due to turbulent blood flow, can increase stress on the plaque, potentially leading to its rupture^7^. All these factors entered in our propensity score and Cox regression analysis, as reported in Table 2, but only CEUS-detected neovascularization was independently associated with TIA/stroke recurrence (hazard ratio, 5.37; 95% confidence interval, 1.36–2.31). Identifying carotid plaque instability is important because it indicates an increased risk of stroke or other cardiovascular events. Imaging techniques such as ultrasound, MRA or CTA can help assess plaque stability by evaluating plaque composition, size, and characteristics^8^. Management of carotid plaque instability typically involves aggressive risk factor modification, including lifestyle changes (such as diet, exercise, and smoking cessation) and medications such as statins, single (or double antiplatelet agents), and/or anticoagulants to stabilize the plaque and reduce the risk of thrombosis and/or embolism. In some cases, surgical intervention, such as carotid endarterectomy or carotid artery stenting, may be considered to remove or stabilize the unstable plaque and restore proper blood flow. Atherosclerotic plaque vascularization, also known as neovascularization, refers to the formation of new blood vessels within or around an atherosclerotic plaque. This process occurs in response to the inadequate blood supply to the growing plaque and the hypoxic conditions within it. Neovascularization is the body’s attempt to supply oxygen and nutrients to the hypoxic areas within or around the plaque. However, the newly formed blood vessels are often fragile and leaky, leading to increased inflammation and plaque instability. Neovascularization is commonly observed in advanced atherosclerosis and is associated with vulnerable plaques that have a higher risk of rupture and causing cardiovascular events such as heart attack or stroke. Detecting neovascularization within atherosclerotic plaques can be challenging but can be achieved through advanced imaging techniques such as contrast-enhanced ultrasound, MRA or CTA^9^. These imaging examinations are expensive and time consuming. In case of CTA, it exposes patients to X ray, on the other side, MRA identifying neovascularization within plaques may help in risk assessment and guiding treatment decisions in individuals with advanced atherosclerosis but it is not adoptable for each patient (e.g. claustrophobia, intra-corporeal metal splinters, intra-corporeal device not MR compatible). Transient ischemic attack (TIA) can occur in individuals with low-grade carotid artery stenosis, although the risk is generally lower compared to cases of more severe stenosis. In low-grade carotid artery stenosis, the narrowing of the artery is typically less than 50%^10^. It is essential to recognize that even low-grade stenosis can contribute to TIA under certain circumstances, first of all plaque vulnerability^11^. In our study, presence of minimal microbubbles which are confined to peri adventitial zone and microbubbles throughout the plaque core are predictive risk factors for TIA/stroke. The absence of microbubble was not related to TIA/stroke recurrence and the best medical therapy (statin, antihypertensive drugs, and single antiplatelet) was enough efficient to prevent further neurological episode overtime^12^.

## Conclusions

Plaque neovascularization, detected by CEUS, is an independent predictor of TIA/stroke recurrence at six-month follow-up in patients with carotid atherosclerosis despite of low-grade stenosis. This finding may guide in the decision between medical therapy and surgical/interventional treatment in the decision making and management of low-grade carotid artery stenosis which is symptomatic for single episode of TIA/stroke. These preliminary data need to be confirmed by large sample of patients and a more longer follow-up period.

## Data Availability

The collected data are available for further analysis and FDA is the responsible for data custody. The study compliances with ethical standards. Each patient signed the specific informed consent to be enrolled in this research and authorized to publish the data in anonymous mode for search purpose only.

